# Just Say No to Postmortem Drug Dose Calculations

**DOI:** 10.1101/2021.04.13.21255383

**Authors:** Peter D Maskell

## Abstract

It has long been known that forensic and clinical toxicologists should not determine the dose of a drug administered based on postmortem blood drug concentrations but to date there has been limited information as to how unreliable these dose calculations can be. Using amitriptyline as a model drug this study used the empirically determined pharmacokinetic variables for amitriptyline from clinical studies and clinical, overdose (where the individual survived) and death (ascribed to amitriptyline toxicity) case studies in which the dose of drug administered or taken was known. Using these data, standard pharmacokinetic equations and general error propagation it was possible to estimate the accuracy of the consumed dose of amitriptyline compared to the actual dose consumed. As was expected in postmortem cases, depending on the pharmacokinetic equation used, the accuracy (mean +128 to +2347 %) and precision (SD ± 383 to 3698%) were too large to allow reliable estimation of the dose of drug taken or administered prior to death based on postmortem blood drug concentrations. This work again reinforces that dose calculations from postmortem blood drug concentrations should not be carried out.

## Introduction

It is common for forensic or clinical toxicologists to be asked by lawyers or law enforcement personnel to calculate the dose or amount of drug that may have been taken by or administered to a deceased person. The forensic toxicology community have long known that the pharmacokinetic equations used to calculate dose in life from measured drug blood concentrations are unsuitable for calculation of drug doses in death cases (1). Thus guidelines from professional organisations such as the Society of Forensic Toxicologists (SOFT) (2), AAFS standards board (ASB) (3) and United Kingdom Association of Forensic Toxicologists (UKIAFT) (4) recommend that these calculations are not attempted. To date however there has been no published proof as to how unreliable these calculations can actually be. This study estimated the accuracy and precision that can arise in postmortem pharmacokinetic dose calculations using an example drug (amitriptyline) published clinical and case study data and general error propagation methodology (5). This work was also able to determine the contribution of each of the variables to the overall uncertainty of the estimate of the dose taken (or administered).

### Standard Pharmacokinetic Equations

The most common pharmacokinetic equation for the calculation of the dose (at equilibrium and assuming complete absorption of the drug) is: -

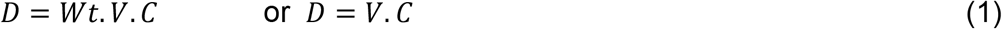

D = dose of drug (at the time of measurement of plasma/blood drug concentration) (mg), Wt = body weight (kg), C = drug plasma/blood concentration (mg/L) and V = volume of distribution (L/kg) or volume of distribution (L). Although this calculation will only give the dose in the body at the time of measurement. It will not give the dose of the drug administered.

### Calculation of Dose after Intravenous Administration

In order to determine the dose of the drug administered the time elapsed since the dose was administered (t) and the half-life (t½) of the drug would also need to be known. For an intravenous bolus dose of the drug, the equation would be: -

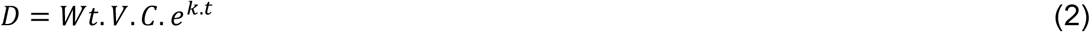

k = elimination rate constant (h^-1^), 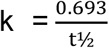, t½ = half-life (h), t = time since administration of dose (h).

This equation however assumes that the pharmacokinetics of the drug fit a one compartment model. In the one compartment model the drug is assumed to be instantaneously distributed throughout the body into a single theoretical compartment. It is more common for the pharmacokinetics of a drug to fit a two-compartment model where there is an initial (α) distribution of the drug from the “first” theoretical compartment (commonly considered to be the circulation) to a “second” theoretical compartment (commonly considered to be the tissues). The α distribution phase is followed by a second β phase where there is equilibrium between the two compartments and elimination is the predominant factor.

The dose calculation for a drug administered by an intravenous bolus that fits a two-compartment pharmacokinetic model is: -

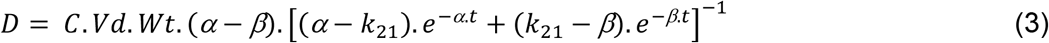

When the drug is infused, a revised equation is used:

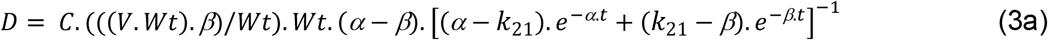

α = elimination rate constant of “distributive” alpha phase (h^-1^)

β = elimination rate constant of “elimination” beta phase (h^-1^) equivalent to k in equation 1.

K_21_ = first-order transfer rate constant from the peripheral compartment to the central compartment (h^-1^)

### Calculation of dose administered after oral administration

The dose calculation is different if the drug is administered (or taken) orally as in this calculation the bioavailability (the amount of drug administered that reaches the systemic circulation) needs to be taken into account. Assuming the drug has been completely absorbed the equation is: -

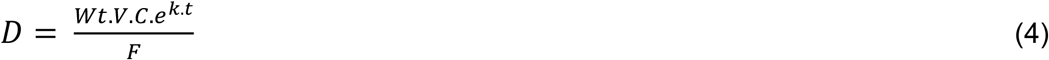

F= Bioavailability (no units)

Again, the complexity is increased if absorption is not complete in which case the rate constant for absorption (Ka) (h^-1^) is needed giving the equation: -

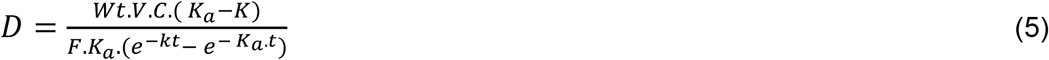

### Determining the precision and accuracy of postmortem dose calculations

In order to investigate the accuracy and precision of the estimation of the dose of a drug taken (or administered) based on a postmortem blood concentration an example drug amitriptyline (a tricyclic antidepressant) was selected. Amitriptyline is a drug for which there is a large amount of clinical data on both the pharmacokinetic parameters but also case studies of doses that were taken in overdoses or deaths attributed to amitriptyline along with the relevant blood amitriptyline concentrations.

### Pharmacokinetic Variables of Amitriptyline

A literature search was carried out to identify studies with empirically determined pharmacokinetic parameters of amitriptyline using www.pubmed.gov and www.scopus.com (06FEB21 and 07FEB21). Any relevant references found in the articles found during the search were also included in the study. Only papers that contained empirically determined amitriptyline pharmacokinetic parameters from human subjects listed in equation 1 - 5 were included. The data obtained from the publications was compiled in Microsoft Excel 2015 (Microsoft Corporation. Redmond. USA). In order to determine if the pharmacokinetic data collected from previously published studies was normally distributed a D’Agostino & Pearson omnibus normality test was carried out using GraphPad Prism version 6.01 for Windows (GraphPad Software, La Jolla California USA, www.graphpad.com) a p value of ≤ 0.05 was considered significant. Only the half-life (t½) and its linked variable the elimination rate constant (β) were found not to be normally distributed (data not shown). Table 1 lists the mean, mode, standard deviation (SD), range and coefficient of variance (%CV) that were used for the “average” individual for the calculation of uncertainty and accuracy of the administered dose using equations 1 - 5. In one study (Burch and Hullin (6)) the raw data of the blood concentration versus time was available and these data were used to determine the variables required for equations 1 - 5 using the Excel plugin PKsolver 2.0 (7).

**Table 1:** Mean Pharmacokinetic Variables for Amitriptyline from the clinical studies. Medians are also given for variables that are not normally distributed.

| Variable | Mean Value | Uncertainty (S.D.) | %CV | Median | Range (Min – Max) | Number of Subjects | Ref |
| --- | --- | --- | --- | --- | --- | --- | --- |
| $\alpha$ ($\text{h}^{-1}$) (normal) | 3.97 | 1.16 | 29.24 | | 2.98 – 5.62 | 4 | (13) |
| $\alpha$ ($\text{h}^{-1}$) (in overdose) | 0.364 | 0.079 | 21.78 | | 0.224 – 0.462 | 7 | (29) |
| $\beta$ ($\text{h}^{-1}$)/$K$ ($\text{h}^{-1}$) (normal) | 0.046 | 0.031 | 67.64 | 0.0370 | 0.015 – 0.230 | 76 | (6, 13, 30–37) |
| $\beta$ ($\text{h}^{-1}$)/$K$ ($\text{h}^{-1}$) (in overdose) | 0.023 | 0.014 | 59.26 | | 0.008 – 0.059 | 21 | (29, 38, 39) |
| $K_{21}$ ($\text{h}^{-1}$) | 0.287 | 0.067 | 23.41 | | 0.211 – 0.345 | 4 | (13) |
| $V$ (L/Kg) | 22.77 | 11.57 | 50.81 | | 6.40 – 45.50 | 33 | (13, 32–35) |
| $V$ (L) | 1581 | 852 | 53.93 | | 459 – 3325 | 29 | (13, 32, 34, 35) |
| $F$ (no units) | 44.1 | 9.51 | 21.57 | | 31.00 – 62.00 | 21 | (32–34) |
| $K_a$ ($\text{h}^{-1}$) | 1.60 | 0.88 | 54.88 | | 0.50 – 3.40 | 26 | (30, 36) |
| $T_{1/2}$ (h) | 18.82 | 7.67 | 40.76 | 18.76 | 3.51 – 47.14 | 76 | (6, 13, 30–37) |
| Wt (kg) | - | 0.4 | - |  | n/a | n/a | (40) |
| $C$ (mg/L) | <0.1 mg/L<br>0.1 to 10 mg/L<br>>10 mg/L | -<br>-<br>- | 12.50<br>10.00<br>7.50 | | n/a | n/a | (41) |

### Uncertainty in dose calculations

The aim of this study was to a) estimate the overall precision of the calculation of dose from a postmortem blood drug concentration using the various relevant pharmacological equations. b) estimate the accuracy of the calculations of dose using pharmacokinetic equations and c) estimate the contribution of each of the variables in the pharmacokinetic equations to the overall uncertainty of the calculation of dose from postmortem blood drug concentrations. This was completed using a) average (mean) pharmacokinetic parameter data from primary literature sources (see Table 1); b) from pharmacological studies where the dose administered was known and c) from case studies (of both overdoses (where the individual survived) and cases where death (which attributed to amitriptyline overdose)). For these case studies to be included in this investigation it was important that the study included a reasonable estimate of the dose of amitriptyline than was taken (see Tables 2 and 3).

**Table 2:** Collation of Amitriptyline fatal overdose case studies in which the dose taken was known and death was attributed to amitriptyline overdose,

| Subject Number | Estimated amitriptyline dose taken (mg) | Subject Weight (kg) | Amitriptyline Concentration at autopsy (mg/L) | Blood Sample Site | Amitriptyline: nortriptyline Ratio | Estimated phase (absorption/elimination) | Time since administration (h) | Reference |
| --- | --- | --- | --- | --- | --- | --- | --- | --- |
| 1 | 2000 | Not given | 0.82 | Not given | 0.72 | elimination | Not given | (42) |
| 2 | 2500 | 67 | 6.00 | Not given | 1.2 | elimination | Not given | (43) |
| 3 | ≤ 2500 | 54 | 18.00 | Not given | ∞ | absorption | Not given | (43) |
| 4 | ≤ 5000 | 55 | 3.00 | Not given | 1.5 | elimination | Not given | (43) |
| 5 | ≤ 2500 | 55 | 5.00 | Not given | ∞ | absorption | Not given | (43) |
| 6 | ≤ 2500 | 70 | 9.00 | Not given | 1 | elimination | Not given | (43) |
| 7 | ≤ 1250 | 57 | 9.00 | Not given | 0.5 | elimination | Not given | (43) |
| 8 | 2000 | Not given | 3.00 | Not given | 3 | absorption | 2 to 4 | (15) |
| 9 | 4500 | Not given | 10.00 | Not given | 3.33 | absorption | 60 to 70 | (15) |
| 10 | 1200 | Not given | 1.00 | Not given | 0.5 | elimination | < 12 | (15) |
| 11 | 7500 | Not given | 3.15 | Not given | 13.1 | absorption | 1 | (14) |
| 12 | 2500 | Not given | 3.00 | Not given | Not given | ? | 7 to 8 | (44) |

**Table 3:** Collation of Amitriptyline non-fatal overdose cases studies in which the dose taken was known.

| Subject Number | Estimated amitriptyline dose taken (mg) | Subject Weight (kg) | Amitriptyline Concentration (mg/L) | Blood Sample Site | Amitriptyline: nortriptyline Ratio | Estimated phase (absorption/elimination) | Time since administration (h) | Reference |
| --- | --- | --- | --- | --- | --- | --- | --- | --- |
| 1 | 2475 | Not given | 0.78 | Serum | 1.01 | elimination | ~ 148 h | (45) |
| 2 | 1875 | Not given | 0.65 | Serum | 1.46 | elimination | 24 | (14) |
| 3 | 3600 | Not given | 0.81 | Serum | 2.52 | absorption | 3 | (14) |
| 4 | 675 | Not given | 0.45 | Serum | 3.03 | absorption | 10 | (14) |
| 5 | 750 | Not given | 0.84 | Serum | 16.20 | absorption | 3 | (14) |
| 6 | 2500 | Not given | 1.45 | Serum | 4.57 | absorption | 11 | (14) |
| 7 | 350 | Not given | 0.49 | Serum | 1.57 | elimination | 2 | (14) |
| 8 | 1500 | Not given | 0.94 | Serum | 5.53 | absorption | 2 | (14) |
| 9 | 375 | Not given | 0.55 | Serum | 5.58 | absorption | 4 | (14) |
| 10 | 1050 | Not given | 0.39 | Serum | 1.66 | elimination | 4 | (14) |
| 11 | 3000 | Not given | 1.28 | Serum | 16.7 | absorption | 1 | (14) |
| 12 | 450 | Not given | 0.46 | Serum | 2.36 | absorption | 2 | (14) |
| 13 | 1750 | Not given | 0.72 | Serum | 5.23 | absorption | 9 | (14) |
| 14 | 2000 | Not given | 0.38 | Serum | 3.31 | absorption | 2 | (14) |
| 15 | 4500 | Not given | 0.50 | Serum | 1.79 | elimination | 3 | (14) |
| 16 | 1500 | Not given | 0.41 | Serum | 0.61 | elimination | 9 | (14) |
| 17 | 3000 | Not given | 1.52 | Serum | 6.49 | absorption | 1 | (14) |
| 18 | 700 | Not given | 0.21 | Serum | 2.30 | absorption | 13 | (14) |
| 19 | 4000 | Not given | 0.77 | Serum | 1.80 | elimination | 15 | (14) |

### Methodology

The accepted method determination of the uncertainty associated with calculations, is that of general error propagation using GUM principles (5) this has a sound mathematical basis and has previously been used forensically for alcohol calculations using the Widmark equation (8–10). Detailed information about this method of error calculation can be found in (11, 12). In order to estimate the total uncertainty associated with the various dose calculation equations and the proportional contribution of each variable to the total uncertainty GUM Workbench EDU Version 2.4.1.384 (Metrodata GmbH, www.metrodata.de) was used. It was assumed that all the variables were independent and that each of the variables were normally distributed. The relevant equations (depending on the case circumstances, clinical; overdose or death) were entered into the software (along with the relevant data from Table 1, 2 or 3). In order to calculate the overall uncertainty of these data and the contribution of each of the variables to the overall uncertainty a randomly selected individual in each case series was selected. For the intravenous administration of amitriptyline (15 mg) subject AJ was used from (13). For the oral administration of amitriptyline (100 mg) subject DB from (6) was used. For the overdose and death cases only, oral administration was investigated as there were only oral overdoses and deaths. Subjects 2 and 3 from study (14) for overdose were used and subjects 9 and 10 from study (15) were used for the death cases. The ratio of amitriptyline to nortriptyline (the major metabolite of amitriptyline) was used to determine if the subject was in the elimination phase or the absorptive phase to allow selection of the relevant pharmacokinetic equation. Based on the work of Bailey & Shaw (16) and Hebb *et al*., (17) this study considered a ratio of amitriptyline to nortriptyline of < 1.5 to be in the elimination phase and > 1.5 to be in the absorptive phase. The accuracy of the calculated dose was determined using the following equation:

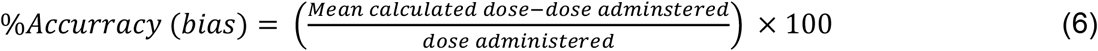

### Accuracy and precision of dose calculations from drug blood concentrations

As can be seen in figure 1, on average for clinical and overdose cases equation 1 was the most accurate and precise although there was a mean underestimation of the dose administered (approx. -0.6 to -82 %). As expected, due to the increased complexity of equations 4 and 5 ((that require additional factors such as half-life (t½), bioavailability (F), time since administration (t) and absorption rate constant (Ka))) on average these equations were less accurate (+23.1 to 549.6%) that equation 1. Again Fig 1. shows that when the pharmacokinetic equations 1, 4 and 5 were used to estimate the dose taken (or administered) in deaths attributed to amitriptyline toxicity there was a very large overestimation of the dose that was taken with an overestimation of between +127.6 % and 2346.0 %. The differences between the actual dose administered compared to the estimated dose administered seen in this study for the clinical group are likely to be due to individual variation in the pharmacokinetic factors as these are known to be person specific. The pharmacokinetic factors are influenced by age, sex, disease state, the physicochemical properties of the specific drug and genetics amongst other factors (for a review see (18)). Even with the most investigated and understood forensically relevant drug, ethanol, there are still large contributing uncertainties from specific pharmacokinetic factors (such as the elimination rate and volume of distribution of ethanol (see reference (19) for summary) and as in the investigation in this work may only be generalised for the “average” person leading to increased uncertainty of the “true” value unless specifically measured in the individual. This something that is impossible in a deceased individual.

**Figure 1:**
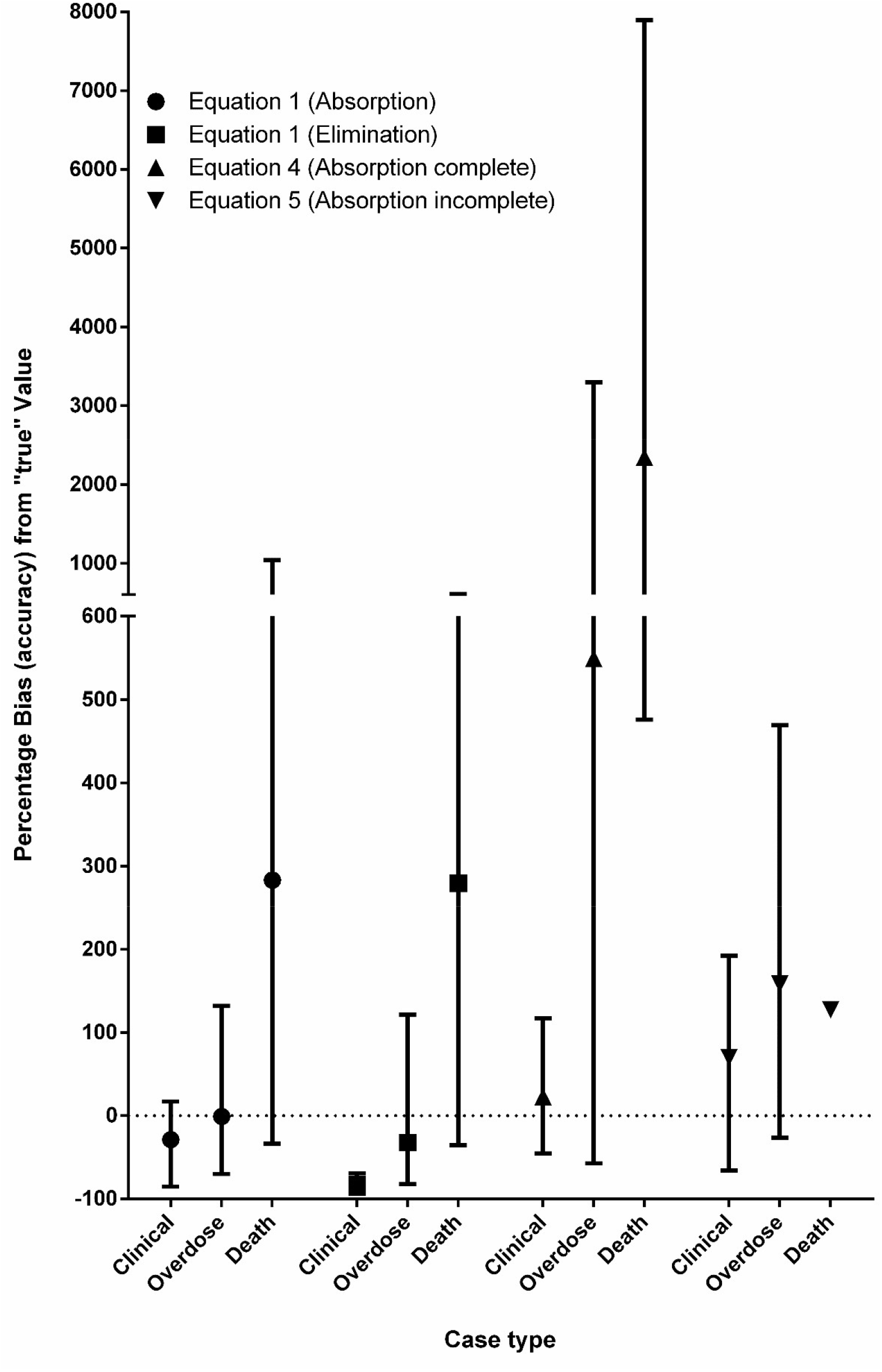
Graph showing the mean percentage accuracy (± SD) of the dose calculation using equations 1, 4 and 5 for 1) Single dose (clinical), 2) Overdose and 3) Death cases. The single dotted line represents the “perfect” dose calculation.

### Uncertainty associated with individual factors of the pharmacokinetic equations

As can be seen in Tables 4 - 7 the equation variables that have the largest influence on the overall uncertainty on the calculation of the dose of a drug taken (or administered) are the elimination rate and the volume of distribution of the individual.

**Table 4:**
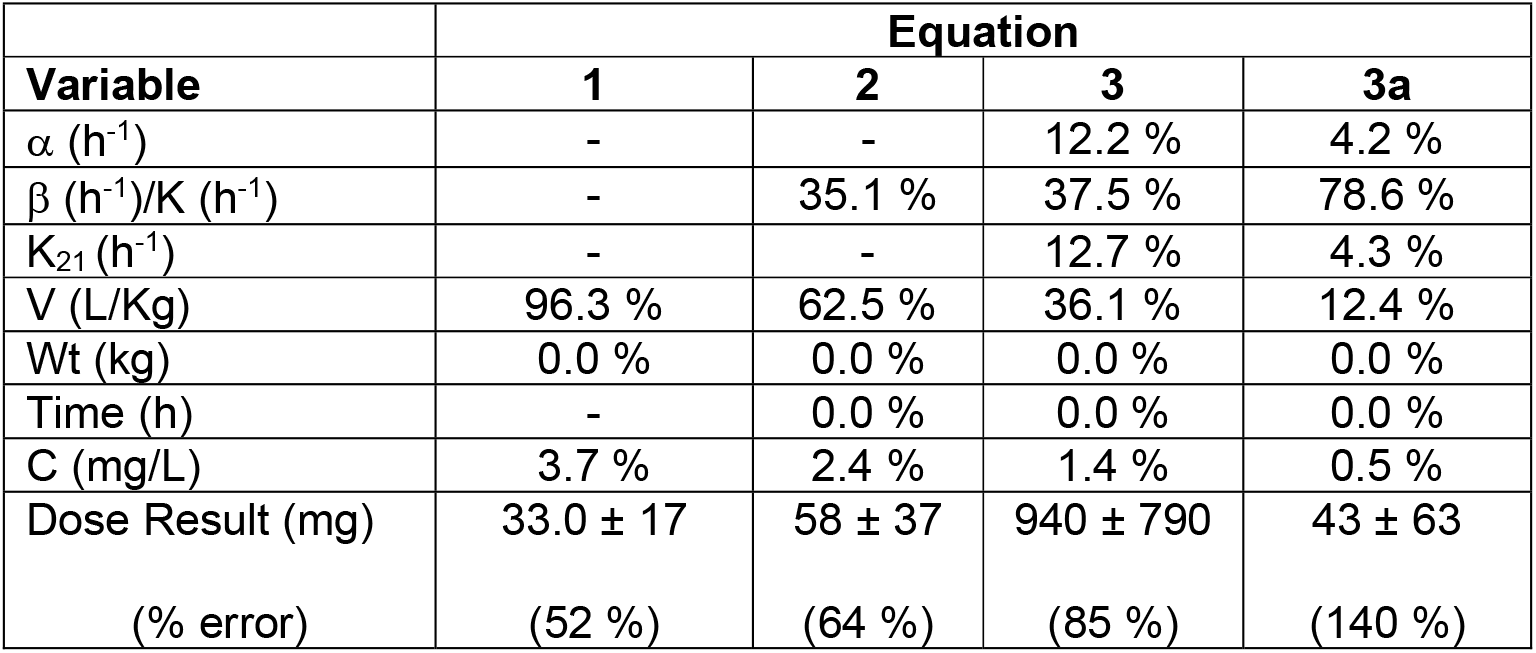
The proportion (as a percentage) that each variable in the dose calculation contributes to the total uncertainty in the various pharmacokinetic equations for clinical cases. Data from subject AJ (13). The blood concentration of amitriptyline was 0.02 ± 0.002 mg/L 12 hours after the dose was administered. The subject had a body mass of 73 ± 0.4 kg. Actual administered dose 15 mg (intravenous infusion). The uncertainty on the calculated dose was expressed as 1 SD.

**Table 5:** The proportion (as a percentage) that each variable in in the dose calculation contributes to the total uncertainty in the various pharmacokinetic equations for clinical cases. Data from subject DB (6). For the absorption the blood concentration of amitriptyline was taken at 3 h (0.058 ± 0.00725 mg/L) for the elimination phase 36 h after dose was administered (0.0087 ± 0.001088 mg/L). The subject weight was not given so volume of distribution (L) used. Administered dose was 100 mg. The uncertainty on the calculated dose was expressed as 1 SD.

| Variable | Equation |  |  |  |
| --- | --- | --- | --- | --- |
|  | 1<br>(absorbing) | 1 (post-absorption) | 4 | 5 |
| F | - | - | 2.3 % | 14.1 % |
| $\beta$ ($\text{h}^{-1}$)/K ($\text{h}^{-1}$) | - | - | 83.9 % | 0.8 % |
| V (L) | 94.9 % | 94.9 % | 13.1 % | 80.1 % |
| $K_a$ ($\text{h}^{-1}$) | - | - | - | 0.7 % |
| Time (h) | - | - | 0.0 % | 0.0 % |
| C (mg/L) | 5.1 % | 5.1 % | 0.7 % | 4.3 % |
| Dose Result (mg) | $92 \pm 51$ | $13.8 \pm 7.6$ | $160 \pm 240$ | $230 \pm 140$ |
| (% error) | (55 %) | (55 %) | (150 %) | (60 %) |

**Table 6:** The proportion (as a percentage) that each variable in in the dose calculation contributes to the total uncertainty in the various pharmacokinetic equations in cases where there was an overdose of amitriptyline. Data for equation 1 (absorbing) and 4 from subject 2 (table 3). Data for equation 1 (post-absorption) and for equation 5 from subject 3 (table 3). The subject weight was not given so volume of distribution (L) used. The uncertainty on the calculated dose was expressed as 1 SD.

| Variable | Equation |  |  |  |
| --- | --- | --- | --- | --- |
|  | 1<br>(absorbing) | 1 (post-absorption) | 4 | 5 |
| $\beta$ ($\text{h}^{-1}$)/$K$ ($\text{h}^{-1}$) | - | - | 25.0 % | 0.3 % |
| V (L) | 96.7 | 96.7 | 62.0 % | 81.6 % |
| F (no units) | - | - | 10.9 % | 14.4 % |
| $K_a$ ($\text{h}^{-1}$) | - | - | - | 0.9 % |
| Time (h) | - | - | 0.0 % | 0.0 % |
| C (mg/L) | 3.3 | 3.3 | 2.1 % | 2.8 % |
| Dose Result (mg) | 1030 ± 560 | 1280 ± 700 | 4000 ± 2800 | 3100 ± 1800 |
| (% error) | (55 %) | (55 %) | (68 %) | (60 %) |

**Table 7:** The proportion (as a percentage) that each variable in in the dose calculation contributes to the total uncertainty in the various pharmacokinetic equations in cases where death was attributed to amitriptyline overdose. Data for equation 1 (absorbing) and 4 from subject 9 (table 2). Data for equation 1 (post-absorption) and for equation 5 from subject 10 (table 2). The subject weight was not given so volume of distribution (L) used. The uncertainty on the calculated dose was expressed as 1 SD.

| Variable | Equation |  |  |  |
| --- | --- | --- | --- | --- |
|  | 1 (absorbing) | 1 (post-absorption) | 4 | 5 |
| $\beta$ ($\text{h}^{-1}$)/K ($\text{h}^{-1}$) | - | - | 76.0 % | 5.7 % |
| V (L) | 96.7 % | 96.7 % | 20.4 % | 77.9 % |
| F (no units) | - | - | 3.6 % | 13.7 % |
| $K_a$ ($\text{h}^{-1}$) | - | - | - | 0.0 % |
| Time (h) | - | - | 0.0 % | 0.0 % |
| C (mg/L) | 3.3 % | 3.3 % | 0.0 % | 2.7 % |
| Dose Result (mg) | 15800 ± 8700 | 1580 ± 870 | 160000 ± 190000 | 4600 ± 2800 |
| (% error) | (55 %) | (55 %) | (120 %) | (61 %) |

This is the same as for ethanol (10) however unlike ethanol where the overall uncertainty for the calculation of results using pharmacokinetic equations is approx. ±20 % (1SD) (8–10) in this study (as seen in Tables 4 - 7) the calculation of the dose of amitriptyline taken has an uncertainty (precision; 1SD) of 52 to 150 % (clinical); 55 to 68 % (overdose) and 55 to 120% (death) a lot larger than those seen with ethanol.

### Postmortem changes and their influence on pharmacokinetic parameters

The changes in the postmortem environment of the body also increase the uncertainty for various of the variable in the pharmacokinetic equations. Postmortem redistribution is the phenomenon known as the “toxicological nightmare” (20). Postmortem redistribution is the change in drug concentration at a specific sampling site after death (20). It is more common for drug concentrations to increase after death at specific sampling sites (sometimes up to 10-fold) (21) but they can also decrease (22), thus there could be a large under (or overestimation) of the drug concentration at the time of death when compared to the measured postmortem drug concentration adding to the uncertainty of the actual blood concentration parameter used in pharmacokinetic equations. Femoral blood is the postmortem sample that is considered to be the “least affected” by postmortem redistribution (23), but recent studies suggest that popliteal blood may be a “better” sample to use than femoral blood(24, 25). However, any postmortem sample is liable to be affected by postmortem redistribution. To date there are no markers to allow determination of the amount of postmortem redistribution that may, or may not have occurred since death and the time the blood is sampled. Thus, the uncertainty of the concentration of the drug will be likely be larger than used in this study (see Table 1). The of half-life and volume of distribution of drugs are also likely be different in fatal drug intoxications compared to those observed in life. For example, the data in Table 1 demonstrates that on average the elimination rate concentration decreases by about half in overdose cases when compared to clinical cases (from 0.046 ± 0.031 h^-1^ to 0.023 ± 0.014 h^-1^) this is likely to decrease further in fatal intoxications as metabolic enzymes and other process become saturated and drug elimination move from 1^st^ order to zero-order elimination. The mean volume of distribution measured in normal individuals is also likely to be different than that found in fatal intoxication cases as in the postmortem environment there are changes in pH (leading to changes in the ionisation of drugs) and drug partitioning (due to loss of cell membrane integrity) along with other changes such as cell lysis (26). Another factor that may be altered in fatal intoxications is that of absorption. The absorption of drugs from the stomach, and thus the bioavailability (F) of a drug, is also assumed to cease in the pharmacokinetic equations. However depending on the case circumstances this has been shown not to be the case especially where tables (or ethanol) reman in the stomach after death (27, 28). The changes of these variables (volume of distribution, elimination rate (half-life), drug concentration and absorption (bioavailability)) in a specific individual leading up to and after death compared to normal healthy individuals is likely to be unknowable. As this study demonstrates these added uncertainties lead to poor accuracy and precision of postmortem dose estimations when compared to the actual doses consumed. The changes in the postmortem environment also make other calculations that are commonly carried out in ethanol forensics such as back-calculations (retrograde extrapolation) highly unreliable. Even in living individuals these calculation are likely to be unreliable due to the lack of large studies of the elimination rate and volume of distribution of specific drugs.

## Conclusions

As expected, both the uncertainty and the accuracy of the calculation of the dose of a drug administered (or taken) from a postmortem drug concentration are too large to allow reliable estimation of the dose of drug taken or administered prior to death. This work again reinforces that dose calculations from postmortem blood drug concentrations should not be carried out.

## Data Availability

Data is available on request.

## Acknowledgements

The author wishes to thank Prof Ian Gent, Dr Ben Jones and Pia Walker for their constructive comments on the manuscript.

